# PHO-Agents: A Large Language Model–Powered Multi-Agent System for Predicting Health Outcomes

**DOI:** 10.64898/2026.06.19.26355815

**Authors:** Daling Shi, Tyler Shugg, Michael T. Eadon, Jing Su, Yijiang Chen, Qianqian Song

## Abstract

**Objective:** Predicting health outcomes from electronic health records (EHRs) is challenging because traditional models rely on structured data and often ignore external medical knowledge. We propose an approach that integrates structured EHR with text-based clinical evidence to improve prediction and interpretability.

**Methods:** We introduce PHO-Agents, a multi-agent system powered by large language models (LLMs) for health outcome prediction. Structured EHR sequences are encoded to produce attention-based representations and initial logits, which are converted into patient summaries by a data agent. A retrieval agent gathers relevant clinical guidelines. Research and practical doctor agents independently assess the patient, and a leader agent synthesizes their analyses. Outputs from the EHR-based model and the LLM agents are fused to generate final predictions and explanation reports. PHO-Agents was evaluated on three real-world cohorts: acute kidney injury (AKI) patients (in-hospital mortality), chronic kidney disease patients (AKI onset within two years), and cancer patients receiving immune checkpoint inhibitors (immune-related adverse events within one year).

**Results:** PHO-Agents outperformed single-agent and multi-agent LLM baselines across all cohorts. In the AKI mortality task, it achieved a PR-AUC of 90.20 ± 2.07, compared with 56.46 ± 2.98 for the best single-agent baseline. Similar gains were observed in the ICI and CKD cohorts. Ablation studies showed that both multi-agent reasoning and logit-level fusion contributed to performance improvements, and case analyses demonstrated clinically consistent explanations.

**Conclusion:** PHO-Agents integrates longitudinal EHR modeling with collaborative LLM reasoning, improving predictive performance, interpretability, and robustness across diverse clinical tasks. This hybrid approach offers a trustworthy strategy for real-world clinical decision support.

## INTRODUCTION

As healthcare systems transition toward digital infrastructures, diverse data sources such as laboratory measurements, medications, diagnoses, clinic notes, and medical images, support a wide range of data-driven applications aimed at improving patient care and health outcome prediction^1-3^. Among these resources, structured electronic health records (EHRs) are collected through standardized clinical workflows, making data reliable and well-suited for large-scale deep learning models^4,5^. Over the past decade, deep learning models have played a major role in leveraging structured EHR data for predictive tasks. Models such as recurrent neural networks, temporal convolutional networks, and transformers can capture complex longitudinal patterns in patient trajectories and achieve strong performance in predicting outcomes such as mortality, hospital readmission, chronic disease onset, and treatment toxicity^3,6-8^. However, most EHR-based predictive models rely on structured inputs and lack access to the broader biomedical knowledge that clinicians use during clinical reasoning and therapeutic decision making^9-11^. Without the ability to incorporate biomedical literature or evidence-based clinical guidelines, these deep learning models typically function as “black boxes” and struggle to provide interpretable justifications for their predictions^12,13^. This creates a gap between algorithmic output and real-world clinical reasoning, ultimately limiting the usefulness of these models in practice, where transparent and explainable decision support is essential for clinical adoption^14,15^.

Recent advances in large language models (LLMs) have created new opportunities for health outcome prediction and improved interpretability through natural-language reasoning^16,17^. Compared with traditional deep learning models, LLMs provide strong natural language reasoning capabilities and can synthesize diverse sources of biomedical knowledge, including scientific literature, disease mechanisms, and medical practical guidelines. In particular, Retrieval-Augmented Generation (RAG) techniques enable LLMs to retrieve relevant medical evidence and incorporate it into the reasoning process, generating explanations that resemble expert clinical interpretation. Early applications in clinical question answering, guidelines interpretation, and medical summarization highlight the potential of LLMs to enhance medical decision support by combining evidence retrieval with advanced reasoning^18-24^. Despite these advantages, single-LLM systems face several critical limitations. They may generate inconsistent predictions for the same patient across different data modalities, struggle to integrate specialized domain knowledge, and often underperform models trained on structured EHR data^3,25^. These limitations can lead to instability, hallucinations, and insufficient grounding in real-world clinical data^26^. To address these issues, recent research has explored multi-agent LLM frameworks that simulate collaboration among specialized clinical experts. In such multi-agent LLM systems, different agents are assigned distinct roles, such as diagnostician, evidence reviewer, or conflict resolver, allowing complex tasks to be decomposed into subtasks handled through collaboration and iterative debate^18,21,24,27-29^. Frameworks such as MedAgents^24^, ReConcile^18^ and Multi-Agent Debate (MAD)^21^ demonstrate that structured coordination across agents reduces reasoning errors, mitigates hallucinations, and improves transparency.

Taken together, these observations highlight a critical gap in current approaches. Structured EHR models provide strong predictive performance by effectively capturing longitudinal patterns in patient data, yet they offer limited interpretability and lack access to the broader biomedical knowledge that underpins clinical decision-making. In contrast, LLM-based approaches provide rich reasoning capabilities and can synthesize diverse sources of biomedical knowledge^12,15^, but they often lack grounding in structured patient data and may produce unstable or inconsistent outputs. The strengths of these paradigms are therefore highly complementary, addressing each otherts key limitations. Integrating these approaches offers a promising opportunity to develop predictive systems that are both accurate and interpretable. Multi-agent LLM frameworks further enable structured collaboration among specialized reasoning components, allowing the system to evaluate evidence, discuss alternative interpretations, and produce more reliable conclusions. These complementary capabilities motivate the development of hybrid architectures that combine structured predictive modeling with collaborative LLM reasoning to support more trustworthy and clinically meaningful health outcome prediction.

To bridge this gap and fully leverage these complementary strengths, we propose PHO-Agents, a hybrid multi-agent framework that integrates structured EHR modeling with collaborative LLM reasoning for predicting health outcomes. The framework combines two important data sources: structured EHR features, and external biomedical knowledge retrieved from PubMed and specialized clinical practice guidelines. PHO-Agents employs a coordinated reasoning process across five specialized agents, i.e. data agent, retrieval agent, research doctor agent, practical doctor agent and leader agent. Within this framework, structured EHR data are first processed by a structured EHR model, and the predictive signals are provided to the multi-agents reasoning module. In this module, specialized agents are responsible for interpreting patient data, retrieving relevant medical evidence, performing clinical reasoning, and synthesizing final predictions. PHO-Agents offers three main contributions. First, it introduces a unified prediction framework that integrates structured EHR modeling with LLM-based clinical reasoning, bridging structured data-driven prediction and knowledge-driven interpretation within a single architecture. Second, the framework demonstrates improved predictive performance. Across multiple real-world clinical datasets, it outperforms models based solely on structured EHR deep learning as well as systems relying exclusively on LLMs, including both single-LLM and multi-agent approaches. Third, the framework demonstrates practical deployability and efficiency. Only the structured predictive model requires training, avoiding LLM fine-tuning and enabling implementation with modest computational resources and low API cost.

## METHODS

### Cohort Definition

This study utilized structured patient-level EHR data from two sources: the MIMIC-IV critical care database^30^ and the University of Florida Health Integrated Data Repository (UFHealth IDR)^31^. MIMIC-IV is a publicly available database developed by the MIT Laboratory for Computational Physiology and contains de-identified electronic health records from patients admitted to the Beth Israel Deaconess Medical Center^30^. The UFHealth IDR is a secure clinical data warehouse and aggregates data from UF Health multiple clinical and administrative platforms, including the Epic electronic health record system. The repository contains over one billion clinical observations covering more than 2 million patients, enabling large-scale studies that support scientific discovery and improvements in patient care and healthcare quality^31^. All analyses were limited to adult patients aged 18 years or older. Observation windows for each cohort were defined based on the index diagnosis or treatment dates described below. Patients were excluded if essential demographic fields or outcome relevant time stamps were missing. This study obtained MIMIC-IV access through PhysioNet credentialing and approval from the UF Institutional Review Board (IRB), IRB202600454.

To evaluate the proposed framework across diverse clinical settings, we constructed three cohorts spanning critical care, chronic disease management, and oncology. Specifically, these include (1) an acute kidney injury (AKI) cohort of ICU patients with in-hospital mortality as the outcome, (2) a chronic kidney disease (CKD) cohort with prediction of future AKI risk, and (3) an immune checkpoint inhibitor (ICI) cohort of cancer patients with prediction of immune-related adverse events (irAEs). All cohorts were defined using standardized diagnosis and treatment codes, clinically relevant features, and well-defined observation windows to ensure consistency and robustness across datasets.

For the AKI cohort, we identified a clinically relevant subset of ICU patients who experienced AKI with tubular necrosis from the MIMIC-IV dataset. The primary outcome was in-hospital mortality, defined as discharge disposition recorded as death. AKI with tubular necrosis was identified using ICD-9 code 584.5 and ICD-10 code N17.0. Patients were required to have at least one ICU stay with complete demographic information, complete admission and discharge time stamps, and sufficient longitudinal clinical measurements. To construct a structured and time ordered analytic dataset, we selected vital signs and laboratory features commonly used in critical care assessment. These included demographics (sex, age, height, and weight), hemodynamic indicators (systolic, diastolic, and mean blood pressure and heart rate), respiratory measurements (respiratory rate, oxygen saturation, and fraction of inspired oxygen), metabolic or laboratory values (blood glucose and pH), body temperature, and neurological assessments captured by the three components of the Glasgow Coma Scale. Record time was retained to preserve longitudinal ordering, and a binary outcome label was included for conducting supervised learning.

For the CKD cohort, we included patients with a diagnosis of CKD from the UFHealth IDR. The primary outcome was the occurrence of AKI within two years following the initial CKD diagnosis. CKD cases were defined using ICD-10 codes N18.x and ICD-9 codes 585.x, while AKI events were identified using ICD-10 codes N17.x and ICD-9 codes 584.x. Patients with less than two years of follow-up were excluded to ensure adequate outcome observation. Laboratory features were selected based on clinical relevance and prevalence, including renal function indicators (e.g., serum creatinine, blood urea nitrogen), hematologic parameters (e.g., hemoglobin), serum glucose, electrolytes (e.g., sodium, potassium, calcium), liver function markers (e.g., bilirubin, alkaline phosphatase), and differential cell counts. Demographics (age, sex, and race) were also included to provide patient context and support adjustment for potential confounders.

For the ICI cohort, we identified cancer patients who received ICI therapy from the UFHealth IDR. The primary outcome was the occurrence of irAEs within one year after initiation of ICI therapy. Cancer diagnoses were identified using ICD-10 codes C00–C97 and ICD-9 codes 140–209. ICI exposure included FDA-approved PD-L1 inhibitors (e.g., atezolizumab, durvalumab, avelumab), PD-1 inhibitors (e.g., nivolumab, pembrolizumab, cemiplimab), and CTLA-4 inhibitors (e.g., ipilimumab), administered alone or in combination. Exposure was identified using RxNorm, NDC, and HCPCS codes across prescribing, dispensing, administration, and procedure tables. The earliest recorded administration date was defined as the index treatment date. irAEs were defined as new diagnoses occurring within one year after treatment initiation, based on ICD-9 and ICD-10 codes covering multiple organ systems. Patients with missing treatment dates or insufficient follow-up were excluded.

### Data Preprocessing

All three datasets were processed using a unified and standardized preprocessing pipeline to ensure consistency and compatibility with subsequent modeling frameworks. For longitudinal preprocessing, laboratory records were aggregated at the patient level and strictly ordered according to recorded timestamps to preserve clinical progression. To ensure sufficient longitudinal signal for sequence modeling, the analysis was restricted to patients with an adequate number of laboratory visits, with cohort-specific minimum thresholds defined a priori. For patients with extensive records, sequence length was standardized by selecting a fixed number of the most recent visits per patient, truncating backward in time from the latest record. This strategy ensured consistent sequence lengths across patients while prioritizing clinically proximal measurements for prediction. The truncation length was determined separately for each dataset according to sampling density and data availability. Measurement frequency varied substantially across cohorts, with approximately daily sampling in the ICU cohort and irregular, event-driven sampling in the ICI and CKD cohorts. To avoid introducing artificial signals, observations were not resampled or interpolated to fixed intervals. Instead, the original visit structure was preserved, and time intervals were explicitly encoded as model features. For each visit, derived variables included time since the first recorded measurement and the interval since the previous visit, which were incorporated as numeric features to the visit-level laboratory feature.

For cleaning and splitting, missingness patterns were assessed across all features, and variables with excessive proportions of missing values were excluded from further analysis. Outliers were identified using modified interquartile range (IQR) based thresholds to minimize removal of clinically plausible extreme values. Values flagged as outliers were treated as missing and subsequently addressed during imputation. Within each patientts longitudinal sequence, missing values were imputed using a last observation carried forward strategy^32^, whereby the most recently recorded measurement was carried forward to replace subsequent missing values, consistent with clinical practice. For features with missing values at the initial time point, imputation was performed using the median value computed from the training set. All continuous variables were normalized using z-score scaling. The normalization parameters, including feature-wise means and standard deviations, were estimated exclusively from the training set and subsequently applied to the validation and test sets to prevent data leakage and preserve strict separation between model development and evaluation phases. Each dataset was independently partitioned into training, validation, and test subsets using patient-level stratified sampling according to outcome labels, ensuring consistent class distributions across subsets while preventing information leakage.

After data processing, three datasets consisted of longitudinal patient sequences ordered by laboratory measurement timestamps, with fully normalized and imputed features. These sequences are then converted into a formatted input, including ordered visit-level vectors, timestamp encoding, and sequence masks. This unified structure ensures that all datasets can be modeled consistently and integrated seamlessly into the PHO-Agents pipeline. A summary of dataset characteristics, including sample size, class imbalance, and the proportions of the training, validation, and test sets, is presented in **Table 1** to provide a clear overview of the three cohorts.

**Table 1:**
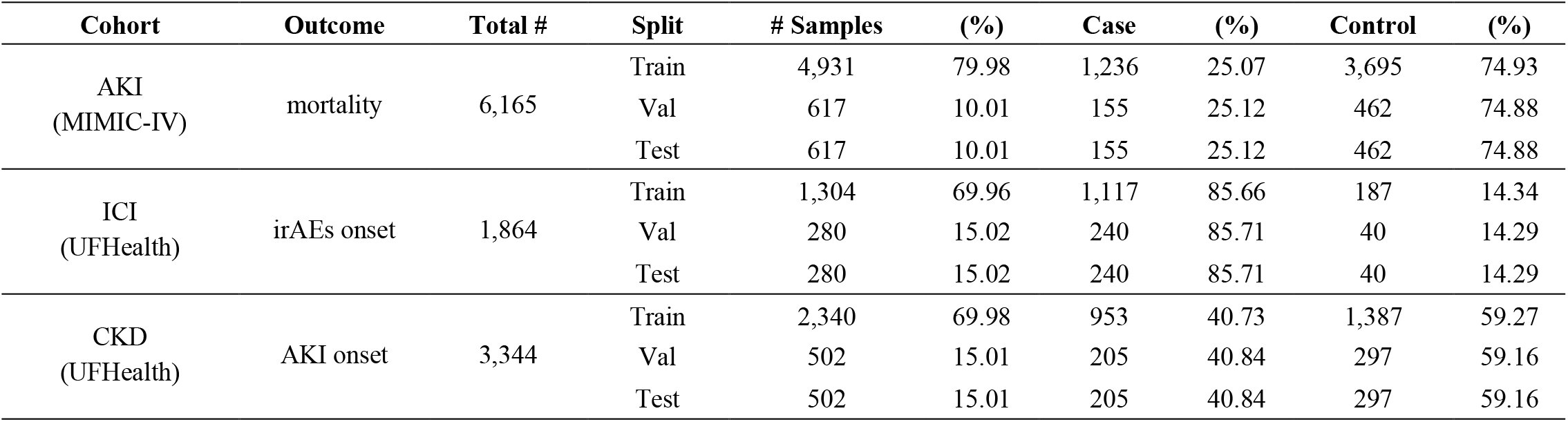
Dataset characteristics, data splits, and outcome distributions. The AKI cohort from MIMIC-IV is used to predict in-hospital mortality, where cases correspond to patient death and controls to survival. The ICI cohort from UFHealth IDR focuses on predicting immune-related adverse events (irAEs), with cases indicating patients who developed irAEs and controls indicating those who did not. The CKD cohort from UFHealth IDR is used to predict AKI onset, where cases represent patients who developed AKI and controls represent those who did not.

To further ensure a rigorous evaluation pipeline, each subset served a strictly defined role throughout the entire PHO-Agents workflow. The training set was used exclusively for EHR-based model training. The validation set was used for early stopping and subsequently for training the fusion module, which integrates predictive logits generated independently by the EHR-based model and the multi-agent reasoning. The test set was reserved solely for final performance evaluation and was not involved in any stage of model development or selection. The multi-agent reasoning module does not involve any learnable parameters and was applied to the validation and test sets independently without any parameter updates, ensuring that test set predictions remain fully uncontaminated. This three-stage separation guarantees that the fusion module is trained on data not used during the EHR-based model optimization, and that all reported test set results reflect unbiased generalization performance.

### Medical Corpus

The research paper corpus was constructed using PubMed, a publicly accessible biomedical literature database maintained by the U.S. National Library of Medicine^33^. To assemble a task-specific literature corpus, structured keyword searches were performed by combining title and abstract terms with MeSH descriptors to improve retrieval precision. Search strategies were tailored to each prediction task to capture key dimensions of relevant medical knowledge, including clinical outcomes (e.g., irAEs, AKI), target populations (e.g., ICU, cancer, or CKD patients), therapeutic contexts (e.g., ICIs), and relevant biomarkers or laboratory measurements. Boolean queries were constructed using both controlled vocabulary and free-text terms, with task-specific adaptations to reflect differences in cohorts, outcomes, and interventions. For example, searches for the ICI cohort emphasized irAEs and treatment-specific terminology, whereas AKI-related searches focused on kidney injury phenotypes and mortality outcomes, and CKD-related searches targeted disease progression and AKI risk. Retrieved articles were ranked by relevance and screened by domain experts, and the titles and abstracts of selected studies were aggregated to form the final corpus used in the retrieval-augmented generation process.

The clinical guideline corpus was constructed from authoritative, evidence-based practice guidelines to provide standardized recommendations for diagnosis, management, and risk stratification. Three primary sources were used: the KDIGO Clinical Practice Guidelines for Acute Kidney Injury^34^, the ASCO Clinical Practice Guidelines for the Management of Immune-Related Adverse Events in Patients Treated With Immune Checkpoint Inhibitor Therapy^35^, and the Merck Manual Professional Version^36^. These sources were selected for their clinical relevance, rigorous evidence synthesis, and widespread adoption. KDIGO guidelines supported AKI-related tasks, including both AKI and CKD cohorts, by providing definitions, staging criteria, risk factors, and management recommendations. The ASCO guidelines were used for the ICI cohort, offering detailed protocols for identifying, grading, and managing irAEs. The Merck Manual served as a general reference, providing complementary clinical knowledge on diseases, symptoms, and laboratory interpretation across conditions.

### System Workflow

PHO-Agents is a hybrid clinical decision-support system that combines the EHR-based predictive model with LLM-driven clinical reasoning (**Figure 1**). By progressively integrating structured patient data with external biomedical knowledge, the system generates risk estimates that are both evidence-based and clinically aligned, supporting transparent and precise decision-making for patient care.

**Figure 1:**
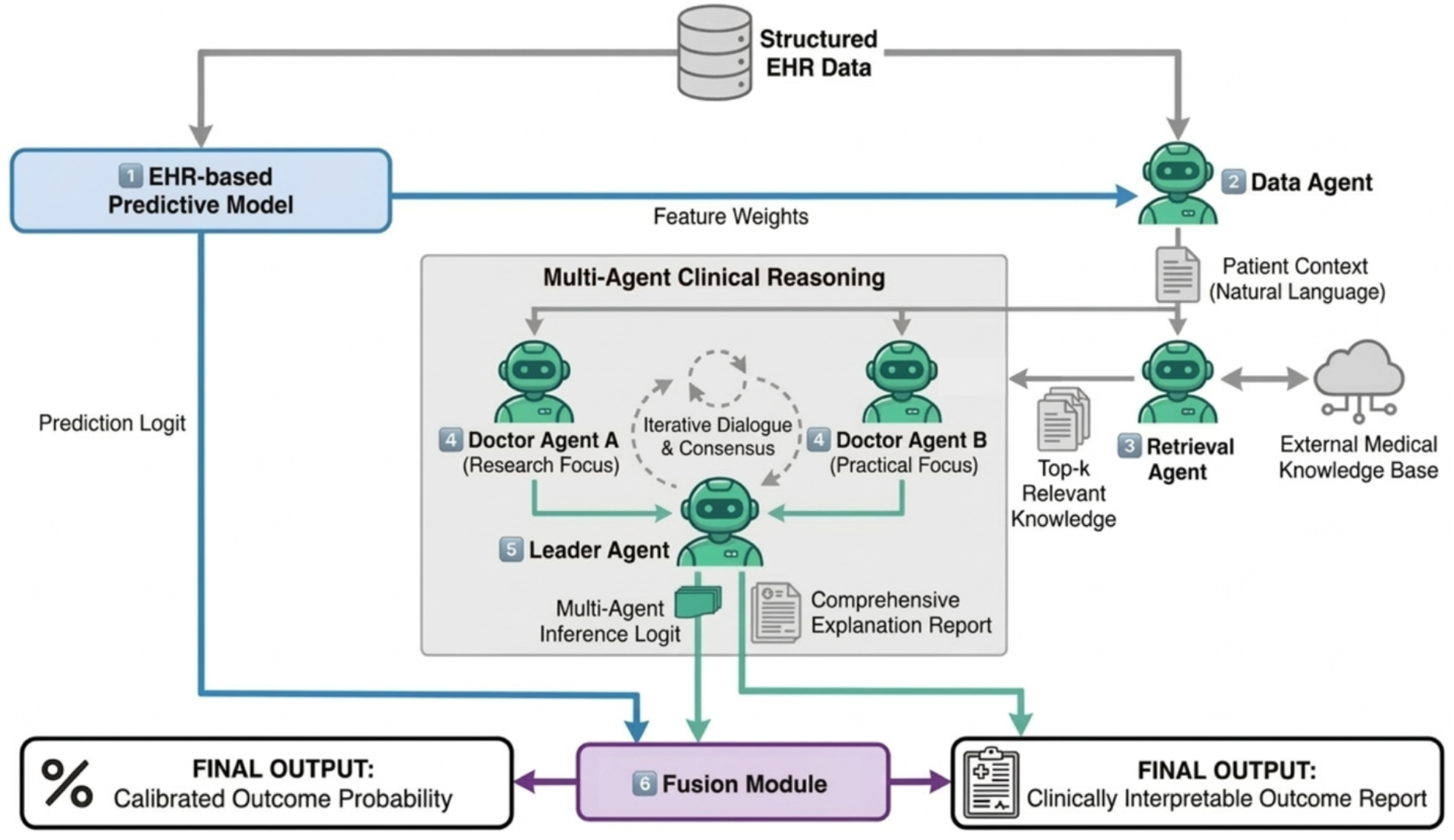
The Framework of PHO-Agents. PHO-Agents integrates the EHR-based predictive model with multi-agent clinical reasoning and knowledge retrieval to generate calibrated, interpretable outcomes. The six-stage pipeline comprises model-based risk estimation, data-to-text context generation, knowledge retrieval, collaborative reasoning among doctor agents, leader-agent synthesis, and fusion of predictions with explanations.

#### Step 1 EHR-based Predictive Modeling

In this study, we adopt an EHR-based predictive model *f*_*BASE*_ with strong performance on longitudinal visit data. Given time-series structured data *X* = {*x*_1_, *x*_2_, …, *x*_t_} where *x*_t_ represents features at time step *t*, the EHR-based model computes and outputs a predictive logit z_RT_:

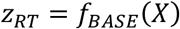

To explain the modelts prediction, we compute and extract SHAP values for each feature, then select the top-K most important features F_k_:

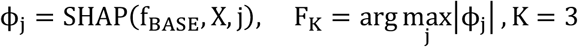

where ϕ_j_ represents feature *j*’s contribution to the prediction.

#### Step 2 Contextual Patient Representation

The data agent transforms the structure patient features and EHR-based model outputs into a templated patient context *C*. This includes demographics, vital signs, lab results, model-derived predictive logits, and feature importance:

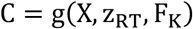

#### Step 3 Retrieval-Augmented Knowledge Injection

To integrate authoritative external evidence, we develop a retrieval agent. This agent is implemented by a dual-encoder system includes two coordinated encoders, the query encoder f_Q_ optimized for patient context and the article encoder f_A_ for longer biomedical documents d_i_, jointly trained in a shared vector space to enable direct semantic similarity computation:

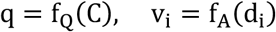

Given the patient context embedding *q*, relevant knowledge D_i_ is retrieved from PubMed abstracts (*P*) or clinical practice guideline repositories (*G*) through inner-product similarity search over a FAISS index of pre-indexed document embeddings:

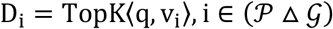

#### *S*tep 4 Agents Clinical Reasoning

Two independent doctor agents, a literature-driven Research Doctor and a guideline-focused Practical Doctor, analyze the patient from complementary perspectives. Each generates a revised logit 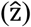, natural-language explanation (*E*), and confidence score (ĉ):

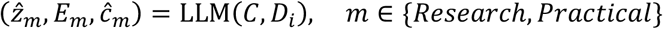

#### *S*tep 5 Leader Agent Coordination

A leader agent orchestrates an iterative consultation process that simulates real-world multidisciplinary case discussions. At each iteration *t*, the leader agent collects the logits and explanations produced by the doctor agents and synthesizes them into a consensus-driven intermediate report:

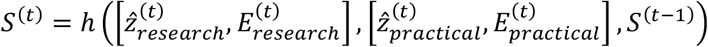

The updated report *S*^(t)^ is redistributed back to both agents to support a new round of reasoning. This iterative process continues until consensus is reached, i.e., the report stabilizes S^*^:

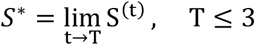

Upon convergence, the leader agent generates a collaborative inference logit z_*L*_ and final consolidated explanation E_L_ summarizing the unified clinical judgment:

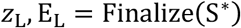

This arbitration mechanism provides a structured approach to consensus building, reduces hallucination risks, and maintains alignment with evidence-backed clinical reasoning throughout the multi-agent interaction.

#### Step 6 Logistic Regression Fusion

To combine the complementary strengths of structured EHR modeling and multi-agent LLM reasoning, we develop a two-stage model stacking framework. First, the EHR-based model provides a baseline predictive logit z_RT_, while the leader agent outputs a knowledge-enhanced estimate logit z_*L*_. These probabilities are then used as features for a logistic regression meta-learner that produces the final prediction:

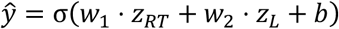

Where σ(·) denotes the sigmoid activation function. Model parameters are learned by minimizing a regularized logistic loss:

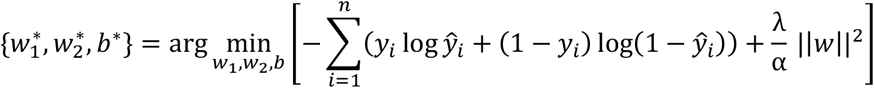

With α selected through cross-validated hyperparameter search. This fusion mechanism adaptively learns the relative reliability of each model and mitigates overfitting through *L*_2_ regularization, thereby improving both predictive accuracy and calibration.

### Implementation Details

In this implementation, the RETAIN model^11^ is selected as the EHR-based model due to its interpretability and strong performance on longitudinal patient visit data. The open-source large language model Llama-3.3-70B-Instruct^37^ served as the primary generative engine and was accessed through the NaviGator API, a secure, institution-managed platform provided by the University of Florida, ensuring that all data processing occurs under strict privacy and security compliance. For embedding and retrieval of medical text, MedCPT^38^ is employed as a dense retrieval model with a bi-encoder architecture that encodes both corpus and queries into shared vector representations. All experiments were conducted on a Linux-based high-performance computing cluster. Jobs were executed on two compute nodes, each equipped with one NVIDIA L4 GPU (24GB VRAM) and 128 GB of system memory, managed by the Slurm workload scheduler. The environment utilized CUDA 12.8. All models were implemented using Python 3.9.23, PyTorch 2.3.0 and Transformers 4.33.2.

### Baselines

To rigorously evaluate the performance of PHO-Agents, we compare against a comprehensive set of state-of-the-art single-agent and multi-agent LLM baselines that span both EHR-driven and knowledge-driven predictive paradigms. Single-agent methods include zero-shot prompting^39^, few-shot prompting^40^ with in-context demonstrations, and self-consistency^41^ decoding, which aggregates multiple reasoning trajectories to produce more stable predictions. These methods assess the intrinsic capability of a standalone LLM to interpret structured clinical data and perform diagnostic reasoning without coordinated knowledge exchange. Beyond single-agent inference, we benchmark against leading multi-agent frameworks designed to enhance medical reasoning through collaborative interactions. ReConcile^18^ employs a round-table consensus mechanism in which multiple LLMs iteratively share explanations and refine predictions based on confidence-weighted evidence. MedAgents^24^ extends this paradigm to healthcare by assigning specialized clinical roles to agents, enabling multidisciplinary reasoning and iterative refinement of decision outputs. Multi-Agent Debate (MAD)^21^ introduces adversarial argumentation, where agents challenge each otherts conclusions under a judgets supervision, promoting diverse perspectives and mitigating degeneration-of-thought issues observed in self-reflective single-agent systems. Collectively, these baselines represent some of the most advanced and widely adopted frameworks for clinical reasoning with LLMs. They provide a strong foundation for evaluating whether PHO-Agentst integration of structured EHR representation learning with coordinated multi-agent medical knowledge exchange offers measurable improvements. Comparisons focus on predictive performance, interpretability, and robustness, which are critical for ensuring reliability and suitability for real-world clinical deployment.

### Evaluation Metrics

All three cohorts exhibit class imbalance, where adverse outcomes such as mortality or AKI onset are minority events. To comprehensively assess predictive performance under this setting, three metrics were adopted: Area Under the Receiver Operating Characteristic Curve (ROC-AUC), Area Under the Precision–Recall Curve (PR-AUC), and the minimum of precision and recall (Min(p, re)). ROC-AUC measures the distinguishing ability of a model across all possible decision thresholds by evaluating its capacity to rank positive cases above negative ones^42^. While ROC-AUC provides an overall estimate of classification separability, it can be overly optimistic for heavily imbalanced datasets. PR-AUC directly focuses on the performance for the positive class by summarizing the trade-off between precision and recall across thresholds^43^. Because it accounts for the prevalence of rare events, PR-AUC is particularly informative for imbalanced clinical outcome prediction tasks, where identifying true high-risk patients is of primary importance. The minimum of precision and recall (Min(p, re)) was used to evaluate the balance between detection accuracy and false-alarm control at a fixed decision threshold. As a conservative metric, Min(p, re) explicitly reflects the weaker component of precision and recall, thereby mitigating the risk of inflated performance driven by optimizing a single metric. These complementary metrics reflect both threshold-independent performance (PR-AUC, ROC-AUC) and threshold-dependent evaluations (Min(p, re)), providing a comprehensive assessment of the modelts predictive performance and practical reliability in imbalanced EHR-based outcome prediction settings.

## RESULTS

### Overview of PHO-Agents

PHO-Agents is a hybrid architecture that combines an EHR-based predictive model with LLM-powered agents to provide interpretable reasoning, including Data Agent, Retrieval Agent, Research Doctor Agent, Practical Doctor Agent and Leader Agent **(Figure 1)**. The PHO-Agents system first applies RETAIN to process structured EHR data, producing predictive logits and feature weights. Then, the Data Agent semantically combines the raw structured patient data with RETAINts outputs and converts them into a natural-language patient context. Using this context, the Retrieval Agent identifies the top-k most relevant documents from the research literature and clinical guidelines to assemble external medical knowledge for downstream reasoning. Utilizing these different knowledge sources, PHO-Agents instantiates two doctor agents with distinct clinical focus areas. Each agent independently evaluates the patient case, generating differential assessments and reasoning chains. A leader agent orchestrates an iterative consultation process that simulates real-world multidisciplinary case discussions: it coordinates dialogue between the doctor agents, synthesizes their perspectives into an evolving summary report, and repeatedly redistributes this report until a stable consensus is achieved. Once consensus is reached, the leader agent produces a collaborative inference logit and an explanatory report. Finally, PHO-Agents integrates the RETAIN-derived logit with the multi-agent inference logit using a logistic-regression fusion module to generate the systemts final prediction. The output includes a calibrated outcome probability and an interpretable, clinically aligned outcome report.

### Performance Comparison of PHO-Agents and Baseline Models

**Table 2** provides a comprehensive comparison of PHO-Agents with the standalone EHR-based predictive model, baseline single-agent, and multi-agent LLM systems across three clinically distinct prediction benchmarks. The three cohorts include critical care (AKI, in-hospital mortality), oncology (ICI, irAEs occurrence within one year), and chronic disease management (CKD, AKI onset within two years), as described in the Cohort Definition section. Model performance was evaluated using PR-AUC, ROC-AUC, and Min(p, re), which together capture discriminative ability and robustness under class imbalanced clinical settings. The inclusion of the standalone EHR-based predictive model as a baseline allows for direct quantification of the incremental benefit introduced by the multi-agent reasoning and logit-level fusion components, isolating the contribution of LLM-based clinical reasoning from the underlying EHR predictive signal.

**Table 2:**
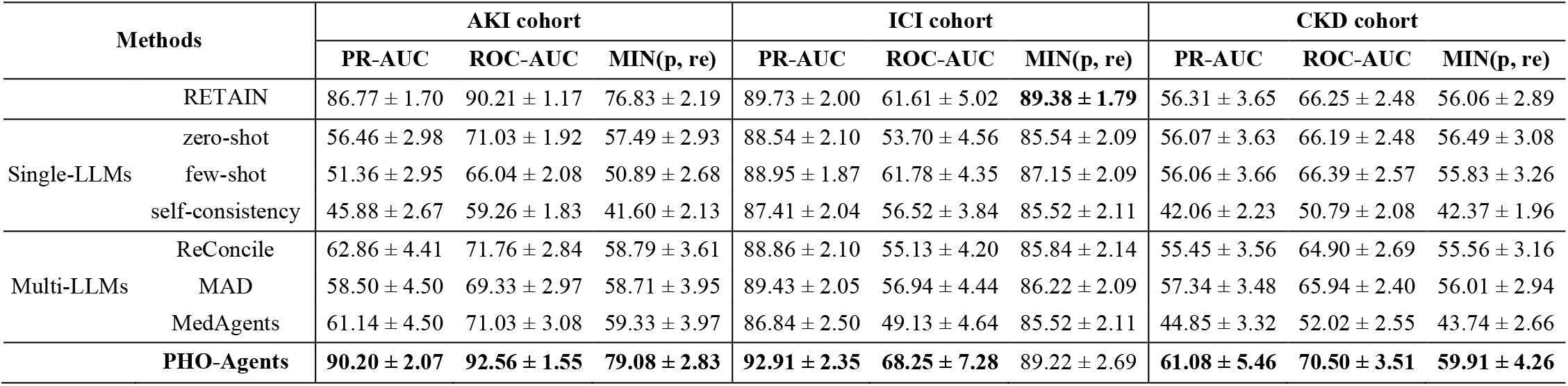
Performance comparison across different cohorts. Results are reported as mean ± standard deviation. Evaluation is conducted on the test sets of the AKI, ICI, and CKD cohorts using PR-AUC, ROC-AUC, and the minimum of precision and recall (MIN(p, re)).

Across all benchmarks, Single-agent LLM approaches, including zero-shot, few-shot, and self-consistency prompting, consistently underperformed the standalone EHR-based model across all cohorts, particularly on PR-AUC. For example, in the AKI mortality task, self-consistency achieved a PR-AUC of only 45.88 ± 2.67, compared with 86.77 ± 1.70 for the EHR-based model alone, highlighting the fundamental limitation of LLM-only approaches when applied to structured clinical prediction tasks without grounding in EHR-derived signals. Multi-agent LLM baselines, including ReConcile, MAD, and MedAgents, showed marginal improvements over single-agent methods, suggesting that structured agent coordination provides some benefit, yet remains insufficient in the absence of quantitative EHR representation. These results underscore that neither single-agent nor multi-agent LLM systems, when operating without structured EHR signals, can match the predictive reliability of data-driven models trained on longitudinal patient records.

PHO-Agents consistently achieved the highest performance across nearly all metrics and cohorts. In the AKI cohort, it attained a PR-AUC of 90.20 ± 2.07 and a ROC-AUC of 92.56 ± 1.55, with MIN(p, re) improving from 76.83 ± 2.19 (EHR-based model alone) to 79.08 ± 2.83, reflecting better precision-recall balance in a critical care setting where false negatives carry significant clinical consequences. In the ICI cohort, PHO-Agents achieved the highest PR-AUC of 92.91 ± 2.35 over the EHR-based model (89.73 ± 2.00) and all LLM-based baselines, highlighting the value of knowledge-grounded reasoning in oncology settings where irAEs prediction requires integrating nuanced immunological evidence beyond structured laboratory data alone. In the CKD cohort, where absolute performance levels were lower across all methods due to the inherent difficulty of predicting AKI onset over a two-year horizon, PHO-Agents still achieved the best performance across all three metrics (PR-AUC: 61.08 ± 5.46; ROC-AUC: 70.50 ± 3.51; MIN(p, re): 59.91 ± 4.26), demonstrating that knowledge-grounded multi-agent reasoning can extract complementary prognostic signals even in tasks primarily driven by routine laboratory measurements. Taken together, these results demonstrate that PHO-Agentst integration of structured EHR representation learning with collaborative, knowledge-grounded multi-agent reasoning yields consistent and robust predictive gains across diverse clinical contexts, with the incremental improvements over the standalone EHR model confirming that LLM-based reasoning contributes meaningfully beyond the EHR signal alone.

### Case Study on Reasoning Capabilities

To further demonstrate the end-to-end collaborative reasoning process of PHO-Agents in a real clinical setting, we present a case study of an older female patient with multi-site cancer undergoing ICI therapy who was assessed as being at high risk for irAEs **(Figure 2)**. The RETAIN model first analyzed the patientts structured longitudinal EHR data, including laboratory measurements such as hemoglobin, blood urea nitrogen, blood glucose, and others, and produced an initial irAEs risk score of 0.86246. In parallel, the model generated feature-level importance weights, identifying Mean Corpuscular Hemoglobin (MCH), Serum/Plasma Creatinine, and AST as the most influential predictors contributing to the risk estimate **(Figure 2, I)**. Based on these structured inputs and model outputs, the data agent translated the patientts numeric clinical profile into a natural-language patient context, summarizing her cancer status, treatment background, abnormal laboratory trends, and model-indicated risk factors. This step provided an interpretable bridge between raw EHR data and downstream multi-agent reasoning.

**Figure 2:**
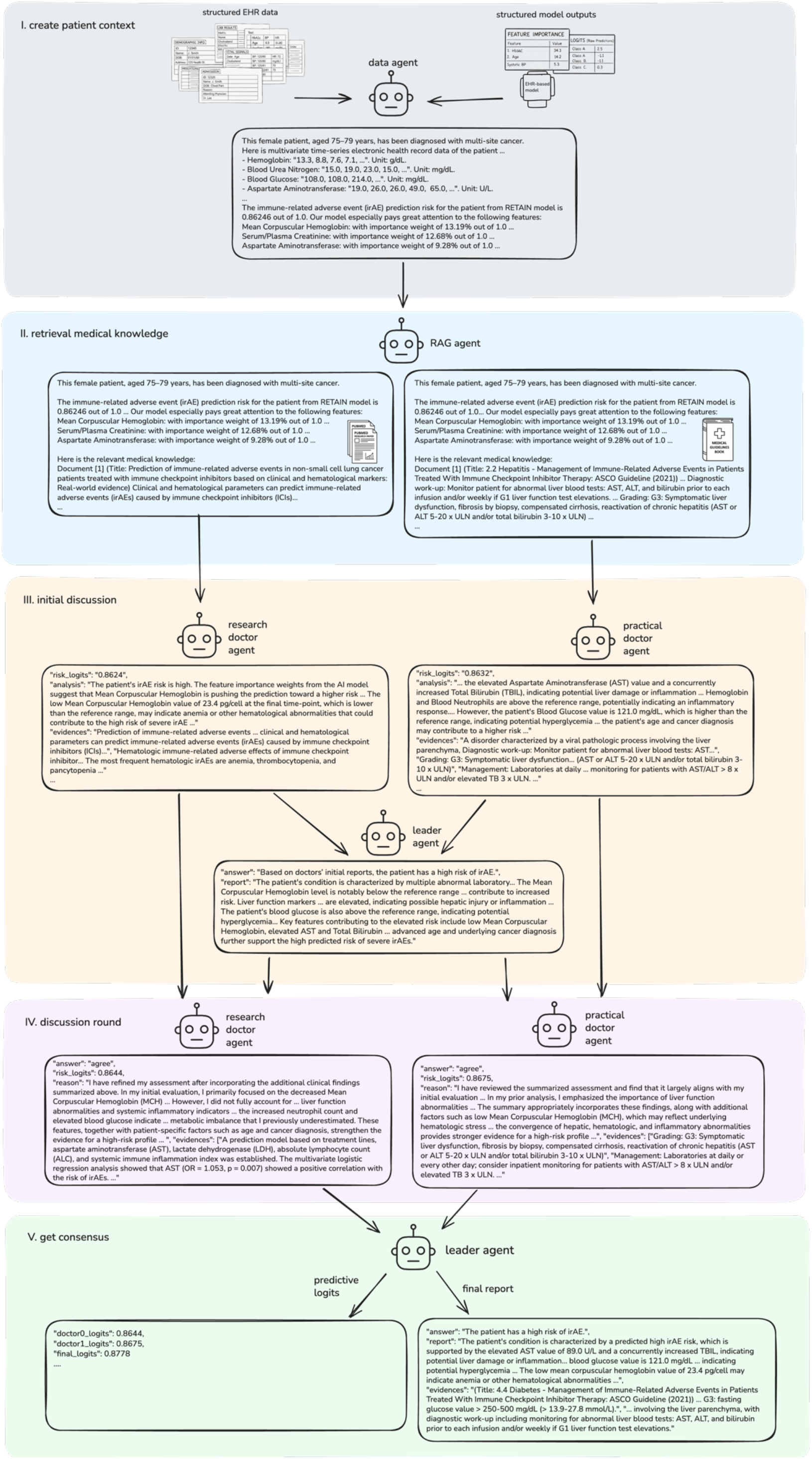
Case study of predicting irAEs in a cancer patient receiving ICI therapy.

Using the generated patient context, the RAG agent retrieved evidence from both peer-reviewed literature and the ASCO Guideline (2021): Management of Immune-Related Adverse Events in Patients Treated With Immune Checkpoint Inhibitor Therapy, with particular emphasis on hepatotoxicity, hematologic abnormalities, and inflammatory responses **(Figure 2, II)**. Two doctor agents then independently interpreted the case from complementary perspectives. The research doctor agent focused more on model-indicated signals and literature-supported associations, emphasizing abnormalities such as reduced MCH and elevated AST as meaningful warning signs linked to irAEs development. The practical doctor agent, from a clinical management perspective, highlighted elevated AST, total bilirubin, blood glucose, and inflammatory-related markers, and connected these findings to possible liver injury, systemic inflammation, and the patientts overall vulnerability given her advanced age and malignancy **(Figure 2, III)**. A leader agent summarized these initial opinions and coordinated a second round of discussion, during which both doctor agents refined their interpretations after reviewing each otherts reasoning and the retrieved evidence **(Figure 2, IV)**. Through this iterative consultation process, the agents reached a stronger consensus that the patient had a high likelihood of irAEs, especially considering abnormal liver function indicators and multiple concurrent laboratory abnormalities. Finally, the leader agent produced a consensus report and integrated the doctor agentst predictive logits with the RETAIN-derived signal to generate the final calibrated prediction, yielding a final risk score of 0.8778 **(Figure 2, V)**. In this case, PHO-Agents not only identified a high-risk patient but also provided a transparent reasoning trail grounded in both patient-specific data and clinical knowledge, illustrating its potential to support interpretable risk assessment in complex oncology settings.

### Ablation Study

To quantify the contribution of collaborative clinical reasoning, an ablation analysis varied the number of doctor agents instantiated within PHO-Agents: (i) 0 agent (only the EHR-based predictive model), (ii) 1 agent, and (iii) 2 agents (full configuration). Performance was evaluated across three clinical prediction tasks: in-hospital mortality prediction (AKI cohort), irAE prediction (ICI cohort), and AKI onset prediction (CKD cohort). As shown in **Table 3**, introducing clinical agent reasoning steadily improves predictive performance compared with the baseline zero-agent configuration. In the AKI cohort, performance exhibited a smaller but stable gain with additional agents. The two-agent setting delivered the best ROC-AUC (92.56 ± 1.55) and improved MIN(p, re) (79.08 ± 2.83), demonstrating that collaborative reasoning can still enrich well-structured clinical domains despite strong baseline signals. In the ICI cohort, where expert reasoning is critical, adding two doctor agents achieved the highest ROC-AUC. Similar performance gains were observed in the PR-AUC metric, which emphasizes the weaker of precision and recall, showing a measurable improvement from 89.73 ± 2.00 (0-agent) to 92.91 ± 2.35 (2-agents). For the CKD cohort, improvements were modest yet directional, with the full 2-agent approach achieving the highest performance. These results indicate that knowledge-based multi-agent assessment can provide useful diagnostic details even in tasks driven by routine laboratory tests and demographic data.

**Table 3:** Ablation study on the number of doctor agents and knowledge sources. We evaluate the impact of varying the number of doctor agents and the inclusion of external medical knowledge from PubMed and clinical practice guidelines across three cohorts: AKI, ICI, and CKD. Results are reported as mean ± standard deviation on the test sets, using AUC-PR, AUC-ROC, and the minimum of precision and recall (MIN(p, re)).

| #Doctor Agents | Corpus |  | AKI cohort |  |  | ICI cohort |  |  | CKD cohort |  |  |
| --- | --- | --- | --- | --- | --- | --- | --- | --- | --- | --- | --- |
|  | PubMed | Practical Guideline | PR-AUC | ROC-AUC | MIN(p, re) | PR-AUC | ROC-AUC | MIN(p, re) | PR-AUC | ROC-AUC | MIN(p, re) |
| 0 | – | – | $86.77 \pm 1.70$ | $90.21 \pm 1.17$ | $76.83 \pm 2.19$ | $89.73 \pm 2.00$ | $61.61 \pm 5.02$ | **$89.38 \pm 1.79$** | $56.31 \pm 3.65$ | $66.25 \pm 2.48$ | $56.06 \pm 2.89$ |
| 1 | ✓ | – | $86.81 \pm 1.71$ | $90.22 \pm 1.19$ | $77.12 \pm 2.07$ | $92.72 \pm 2.32$ | $66.99 \pm 7.22$ | $88.74 \pm 2.36$ | $60.66 \pm 5.56$ | $70.32 \pm 3.58$ | $59.81 \pm 4.29$ |
| 1 | – | ✓ | $86.82 \pm 1.70$ | $90.27 \pm 1.18$ | $76.93 \pm 2.12$ | $92.71 \pm 2.41$ | $67.62 \pm 7.72$ | $88.59 \pm 2.45$ | $60.54 \pm 5.60$ | $70.37 \pm 3.63$ | $59.91 \pm 4.22$ |
| 2 | ✓ | ✓ | **$90.20 \pm 2.07$** | **$92.56 \pm 1.55$** | **$79.08 \pm 2.83$** | **$92.91 \pm 2.35$** | **$68.25 \pm 7.28$** | $89.22 \pm 2.69$ | **$61.08 \pm 5.46$** | **$70.50 \pm 3.51$** | **$59.91 \pm 4.26$** |

To evaluate the contribution of structured and unstructured predictive integration, we conducted an ablation study comparing PHO-Agents with and without the final logistic-regression fusion module. The without fusion condition uses only the multi-agent collaborative inference logit, whereas the full PHO-Agents system incorporates both logits from the EHR-based predictive model and multi-agent reasoning logits into a unified calibrated prediction (**Table 4**). Across all three clinical tasks, fusion consistently improved discrimination performance and class-imbalance robustness. In the AKI dataset, PHO-Agents achieved the strongest PR-AUC (90.20 ± 2.07) and the best MIN(p, re) (79.08 ± 2.83), indicating that integrating structured physiological dynamics with contextual reasoning reduces false-negative risk in critical care settings. Fusion benefits were also demonstrated in the irAE prediction task. The full PHO-Agents pipeline delivered meaningful gains over the without fusion variant in PR-AUC metrics. Additionally, the improvement in the conservative MIN(p, re) metric further demonstrates the role of fusion in achieving balanced clinical trade-offs when identifying high-risk patients for adverse immune responses. While the prediction task in the CKD dataset is primarily driven by structured renal function indicators and comorbidity-related laboratory values, the fusion module still contributed incremental performance gains in both PR-AUC (61.08 ± 5.46) and ROC-AUC (70.50 ± 3.51). These improvements suggest the benefits of integrating the EHR-based predictive model with knowledge-grounded multi-agent reasoning. By combining time-series risk representations with collaborative reasoning, the systemts ability to detect subtle early deterioration signals in CKD patients at risk of progressing to AKI is enhanced. The strengthened MIN(p, re) metric further supports that fusion improves sensitivity to high-risk cases while maintaining precision, which is an essential requirement in proactive AKI prevention and intervention. To summarize, these results underscore fusion as a key mechanism for unifying complementary signal sources, enabling PHO-Agents not only to reason more like clinicians but also to preserve and reinforce quantifiable trends embedded within real-world EHR time-series. The ability to jointly leverage mechanistic risk factors and knowledge-grounded judgment improves robustness and calibration across diverse patient populations.

**Table 4:** Ablation study on logit-level fusion between the EHR-based predictive model and LLM-based inference. We evaluate the effect of fusing the prediction logits produced by RETAIN model with LLM-powered inference across the AKI, ICI, and CKD cohorts. Results are reported as mean ± standard deviation on the test sets using PR-AUC, ROC-AUC, and the minimum of precision and recall (MIN(p, re)).

| Method | AKI cohort |  |  | ICI cohort |  |  | CKD cohort |  |  |
| --- | --- | --- | --- | --- | --- | --- | --- | --- | --- |
|  | PR-AUC | ROC-AUC | MIN(p, re) | PR-AUC | ROC-AUC | MIN(p, re) | PR-AUC | ROC-AUC | MIN(p, re) |
| w/o Fusion | $86.33 \pm 1.76$ | $89.80 \pm 1.23$ | $76.25 \pm 2.24$ | $88.79 \pm 2.24$ | $58.00 \pm 5.00$ | $87.46 \pm 1.96$ | $56.29 \pm 3.55$ | $66.53 \pm 2.44$ | $55.44 \pm 2.88$ |
| PHO-Agents | **$90.20 \pm 2.07$** | **$92.56 \pm 1.55$** | **$79.08 \pm 2.83$** | **$92.91 \pm 2.35$** | **$68.25 \pm 7.28$** | **$89.22 \pm 2.69$** | **$61.08 \pm 5.46$** | **$70.50 \pm 3.51$** | **$59.91 \pm 4.26$** |

### Cost Analysis

To assess real-world deployment feasibility, we quantified the per-patient computational cost of PHO-Agents across the three clinical prediction settings (**Figure 3**). Results show that the system achieves multi-source reasoning with very low latency and minimal operational expense. On average, a full PHO-Agents inference requires only 7 to 9 LLM API interactions per patient, reflecting an efficient consultation process despite multi-agent conversations. The total number of generated tokens remains modest, with per-patient usage below 15k prompt tokens and 2.2k output tokens across datasets. End-to-end inference completes within 1 minute for most cases, indicating near real-time decision support capability suitable for clinical workflow integration. Importantly, the per-patient cost is extremely low, ranging from $0.012 to $0.014, demonstrating that advanced collaborative LLM reasoning can be delivered at a price point competitive with conventional risk prediction models. Notably, the cancer task presents slightly higher API usage, consistent with more extensive knowledge retrieval and longer reasoning traces required for immunotherapy toxicity assessment. Overall, PHO-Agents successfully balances predictive strength, interpretability, and operational efficiency. It enables scalable deployment within cost-constrained healthcare environments.

**Figure 3:**
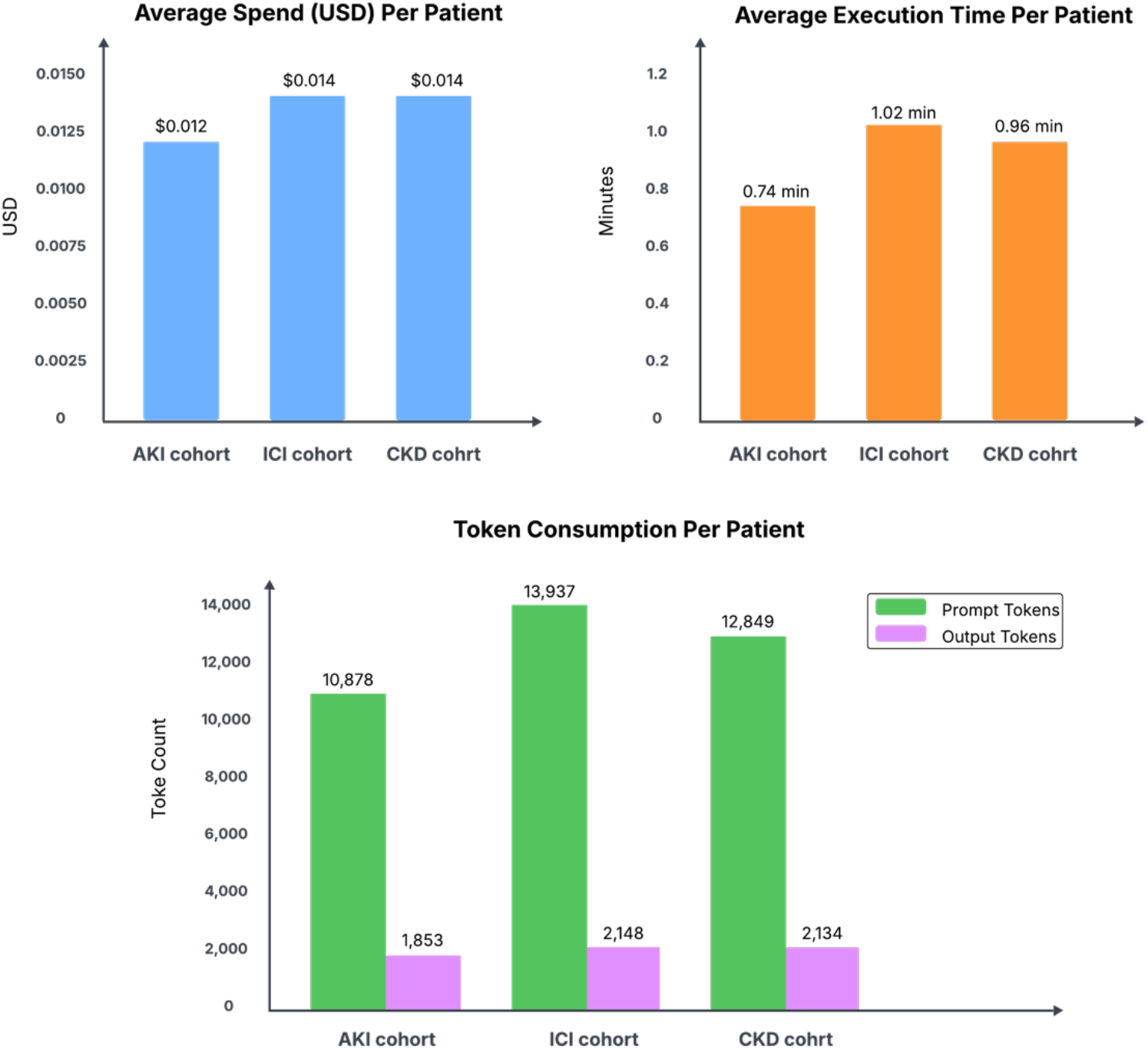
Per-patient computational cost of PHO-Agents across three clinical datasets. We report the average API cost, execution time and number of prompt and output token consumption incurred per patient. Results are shown for the AKI cohort, ICI cohort, and CKD cohort, demonstrating comparable computational overhead and cost efficiency across different disease settings.

## DISCUSSION

In this study, we propose PHO-Agents, a hybrid clinical risk prediction framework that tightly integrates structured EHR time-series modeling with transparent LLM-based multi-agent medical reasoning to advance both predictive performance and interpretability. The system leverages an EHR-based predictive model to capture quantitative disease progression patterns from longitudinal EHR signals, while domain-specialized LLM doctor agents contextualize these signals using medical knowledge grounded in published literature and clinical guidelines. Through coordinated multi-agent debate, revision, and consensus formation, PHO-Agents operationalizes a multidisciplinary consultation paradigm that closely mirrors real-world diagnostic and prognostic workflows. Across diverse disease settings, this synergistic fusion of mechanistic temporal modeling and collaborative knowledge-driven reasoning substantially improves risk stratification performance, yielding superior discrimination and more balanced precision–recall trade-offs compared with single-agent or non-fusion baselines. Beyond accuracy gains, PHO-Agents generates transparent, text-based reasoning chains that articulate how structured clinical trends align with external medical evidence, fostering auditability, clinician trust, and shared decision-making. The iterative consensus mechanism further mitigates individual agent bias and overconfidence, enhancing the robustness and reliability of model outputs. Importantly, these advantages are achieved with minimal operational overhead, maintaining inference costs below $0.02 per patient and average runtimes of approximately one minute, demonstrating that scalable and interpretable multi-agent collaboration is feasible for real-world clinical deployment.

Building on this hybrid design, PHO-Agents exhibits several distinctive advantages that further strengthen both predictive reliability and clinical trustworthiness. First, the framework integrates targeted multi-source medical knowledge systematically. It retrieves and synthesizes evidence from complementary corpora such as PubMed research articles and disease-specific clinical guidelines. Unlike prior multi-agent approaches that often rely on generic web search or single-source prompting^22,28,29^, this knowledge-grounded retrieval strategy supports more contextualized and disease-aware reasoning, improving robustness and generalizability across different medical knowledge domains. Second, PHO-Agents uniquely combines structured EHR representation learning with LLM-based multi-agent interpretability in a tightly coordinated inference pipeline. Structured EHR data are clinically validated and temporally precise indicators of patient physiology. When modeled using deep learning methods, they provide robust quantitative signals for health outcome prediction. Meanwhile, large language models are excellent at contextual interpretation and natural-language reasoning, enabling them to articulate underlying causal patterns and clinical rationale. These two paradigms have complementary strengths: structured data modeling ensures accuracy and numerical fidelity, whereas LLM-based multi-agent reasoning ensures transparency and medical explainability. By coupling both components in a collaborative inference pipeline, PHO-Agents overcomes limitations inherent in single-modality systems and delivers predictions that are both strongly data-driven and interpretable. Finally, PHO-Agents demonstrates robust and consistent performance across diverse disease settings. PHO-Agents consistently improves ROC-AUC, PR-AUC, and balanced precision–recall metrics across three distinct clinical prediction tasks, indicating reliable generalization across varied patient populations and outcome mechanisms. Collectively, these results highlight the adaptability of PHO-Agents for real-world clinical deployment. In such settings, reliable risk estimation, explainable reasoning, and consistency across conditions are prerequisites for clinician adoption and trustworthy decision support^14^.

Despite the demonstrated strengths of PHO-Agents, several important directions remain for future work. First, the current evaluation is limited to retrospective datasets derived from U.S. healthcare institutions. While these settings provide rich and well-curated EHR data, further validation across international cohorts, diverse healthcare systems, and low-resource environments is necessary to establish the frameworkts generalizability and clinical robustness. Prospective studies and external validation on heterogeneous populations will be particularly important for assessing real-world performance and fairness. Second, although PHO-Agents grounds multi-agent reasoning in curated PubMed literature and clinical practice guidelines, the reliability of agent conclusions inevitably depends on the quality, completeness, and timeliness of the retrieved evidence. This dependency is especially pronounced in rapidly evolving clinical domains, where emerging findings may not yet be reflected in established guidelines. Future work could explore adaptive retrieval strategies, continual learning, and evidence quality assessment mechanisms to improve robustness under knowledge uncertainty. In addition, since PHO-Agents relies on LLM-based reasoning, the risk of hallucination cannot be fully eliminated, even when predictive accuracy remains high. Developing safeguards may further enhance trustworthiness. Finally, the current implementation integrates structured laboratory and diagnosis information with textual medical knowledge only. Extending the framework to incorporate additional data modalities is a promising direction for enabling more comprehensive and context-aware clinical reasoning. Together, these directions position PHO-Agents as a step toward scalable, interpretable, and knowledge-aware AI that can meaningfully support precision medicine and trustworthy clinical decision-making.

## Data Availability

All data produced in the present study are available upon reasonable request to the authors.

## Code availability

All code used to develop the PHO-Agents system and generate the results in this study is publicly available under the MIT License at https://github.com/QSong-github/PHO-Agents.

## Author Contributions

D.S. and Q.S. take responsibility for the integrity of the data and the accuracy of the data analysis. D.S., Y.C., and Q.S. contributed to the study concept and design, as well as data acquisition, analysis, and interpretation. D.S., T.S., M.E, J.S., Y.C., and Q.S. drafted the manuscript, performed statistical analysis, and critically revised the manuscript for important intellectual content. Y.C. and Q.S. obtained funding and supervised the study.

## Compliance with Ethics Requirements

This article does not contain any studies with human or animal subjects.

